# Statistical Analysis Plan (SAP): A randomized controlled trial of intensive blood pressure control on cardiovascular risk reduction in patients with atrial fibrillation: the CRAFT trial

**DOI:** 10.1101/2025.06.19.25329849

**Authors:** Xiaolei Lin, Chao Jiang, Zhiyan Wang, Hisatomi Arima, Xia Wang, Laurent Billot, Bruce Neal, Anthony Rodgers, Graham S. Hillis, Anushka Patel, Qiang Li M. Biostat, Jianzeng Dong, Xin Du, Changsheng Ma, Craig S. Anderson

## Abstract

The CRAFT trial is an international, multicenter, randomized controlled, open-label, parallel-group trial with blinded outcome assessment, designed to evaluate whether intensive blood pressure (BP) control can reduce cardiovascular (CV) risks in patients with atrial fibrillation (AF). A total of 1,675 participants with documented AF are randomized to either intensive BP control (target home systolic BP [SBP] <120 mmHg) or standard BP control (target home SBP <135 mmHg). The primary outcome is a hierarchical composite of time to CV death, number and time to first stroke, number and time to first myocardial infarction (MI), and number and time to first heart failure hospitalization (HFH), analyzed using the win-ratio approach. The win ratio and its 95% confidence interval (CI) will be calculated assuming an approximately normal distribution of the log win ratio, with the standard error estimated from the Finkelstein–Schoenfeld Z-statistic. Secondary outcomes include each component of the composite endpoint, all-cause mortality, renal outcomes, quality of life, major bleeding, and other CV events, analyzed using Cox proportional hazards, Poisson, and logistic regression models as appropriate. Sensitivity analyses will use matched win-ratio methods based on CHA[DS[-VASc score components. The planned sample size provides 80% power to detect a win ratio of 1.50 for intensive versus standard BP control after a mean of 3 years of follow-up. Findings from CRAFT will provide robust evidence on optimal BP targets for reducing cardiovascular risk in patients with AF.

**Revison Summary:** This version contains this updated information.

1) We modified the protocol date was corrected to reflect the finalized version.
2) The method for estimating the 95% confidence interval of the win ratio was revised. Instead of using a bootstrap-based approach, the confidence interval is now calculated under the assumption that the log win ratio follows an approximate normal distribution, with its standard error estimated from the Z-statistic of the Finkelstein-Schoenfeld test.
3) The illustrative figure for the win ratio was updated.
4) The definition and analytical approach for subgroup analyses were updated. A detailed definition of ASCVD was added, and the analysis method was revised to use the win ratio stratified for each subgroup instead of including subgroup–treatment interaction terms in the main model.
5) The approach to handling missing data was revised — the previous plan for multiple imputation and sensitivity analyses was removed, and it was specified that no imputation will be conducted.
6) The matched win ratio analysis was updated to specify that matching is performed using the individual components of the CHA[DS[-VASc score and implemented through a nearest neighbour matching method.
7) No statistical test will be conducted among SAE and AESI.

## 2 Introduction

### 2.1 Study synopsis

The Cardiovascular Risk reduction in patients with Atrial Fibrillation Trial (CRAFT) is an investigator-initiated and conducted, international, multicenter, open-label, parallel-group, blinded outcome assessed, randomized controlled trial of intensive blood pressure (BP) control in patients with AF. The aim is to determine whether intensive BP control (target home systolic blood pressure [SBP] <120 mmHg) is superior to standard BP control (target home SBP <135 mmHg) on prevention of the hierarchical composite outcome of time to cardiovascular (CV) death, number of stroke events, time to the first stroke, number of myocardial infarction (MI) events, time to the first MI, number of heart failure (HF) hospitalization events, and time to the first HF hospitalization.

### 2.2 Study population

A total of 1675 patients with documented AF and a standard resting office SBP of 140-179 mmHg (if not on any BP lowering medication) or 125-164 mmHg (if on any antihypertensive medication), are screened for inclusion in an open run-in assessment phase lasting 2 weeks. During the run-in phase, participants have their antihypertensive medication titrated as necessary, to ensure that they have an appropriate stable controlled range of SBP according to measurements of BP taken 3 times in the morning and evening for at least 1 week, with these readings uploaded for central monitoring. Participants with a home SBP 125-154 mmHg during the run-in phase are eligible for inclusion. If the home SBP is ≥155 mmHg or <125 mmHg during the initial run-in phase, the run-in period may be prolonged a further 2 weeks.

### 2.3 Participant eligibility criteria

2.3.1 Inclusion criteria

∘ age ≥18 years;
∘ documented AF, either persistent or paroxysmal (≥2 episodes in the previous 6 months);
∘ home SBP 125-154 mmHg on nil or ≤3 antihypertensive drugs;
∘ presence of at least one of the following CV risk factors: history of thromboembolism (ischemic stroke, transient ischemic attack [TIA], or systemic embolism), diabetes mellitus (DM), coronary artery disease (CAD), peripheral artery disease (PAD), chronic kidney disease (CKD), or an age ≥65 years.

2.3.2 Exclusion criteria

∘ successful AF ablation;
∘ moderate or greater mitral stenosis, or mechanical valve replacement;
∘ home SBP ≥145 mmHg after taking ≥4 antihypertensive medications at full-dose;
∘ inability to upload home BP recordings over ≥5 days in the run-in phase;
∘ secondary hypertension;
∘ SBP <110 mmHg after 1 minute standing;
∘ diagnosis of polycystic kidney disease;
∘ glomerulonephritis treated with immunosuppressant therapy;
∘ an estimated glomerular filtration rate (eGFR) <30 mL/min/1.73m^2^;
∘ any CV event, CV-related procedure, or hospitalization for unstable angina in the past 3 months;
∘ HF with reduced left ventricular ejection fraction (<40%) or clinical severity of grated III-IV in the New York Heart Association (NYHA) classification;
∘ diagnosis of dementia;
∘ pregnancy.

### 2.4 Study interventions

*Intensive BP control group (home SBP <120 mmHg):* most participants in the intensive group are recommended to commence treatment with 2 or 3 antihypertensive agents. If the home SBP remains elevated (ie ≥120 mmHg), another type of antihypertensive drug should be added or the dose of the initial antihypertensive drugs increased. Dosage adjustments or drug additions are made every month until a stable level of home SBP <120 mmHg is achieved. If a participant’s home SBP is >120 mmHg during any 6-month follow-up period, a different antihypertensive medication should be added rather than increasing the dose of a current drug, unless there are contraindications to this approach. If severe symptoms of hypotension should occur, a gradual reduction in medication is recommended.

*Standard BP control group (125 mmHg < home SBP <135 mmHg):* most participants in the standard BP control group are likely to achieve the home SBP target (<135 mmHg) within 3-6 months. The treatment approach aims to achieve a mean home SBP of 130-134 mmHg in as many participants as possible. On the basis of a careful review of prior antihypertensive medications at the randomization visit, investigators are required to determine the best medication strategy in order to maintain a home SBP of 130-134 mmHg, using combination or single drug therapy. Either a dose adjustment or the addition of another drug may be needed if home SBP is ≥135 mmHg during follow-up. Down titration (a reduction of the dose or number of antihypertensive drugs) should be carried out if home SBP is <125 mmHg.

### 2.5 Outcomes

#### 2.5.1 Primary outcome

The primary outcome is a hierarchical composite of time to CV death, number of strokes, time to the first stroke, number of MI events, time to the first MI, number of HF hospitalization events, and time to the first HF hospitalization, compared using the win-ratio approach, after 1600 patients are randomized and who have had a mean of 3 years of follow-up, which is anticipated to be reached by March 2026. Adjudication of the primary outcomes is performed by a blinded and independent Clinical Event Committee (CEC).

Definitions of CV endpoints follow the 2017 Cardiovascular and Stroke Endpoint Definitions for Clinical Trials consensus developed by the Standardized Data Collection for Cardiovascular Trials Initiative (SCTI).^1^

#### 2.5.2 Secondary outcomes

*Each component of the primary composite outcome* - CV death, stroke, MI, HF hospitalization

*Time to the components of the composite outcome* - CV death, stroke, MI,r HF hospitalization All-cause death

*Pathological subtypes of stroke* - ischemic stroke separate to intracerebral hemorrhage

*Renal outcomes -* The time to a first occurrence of CKD progression or incident albuminuria, measured at baseline, 6 months, and every year during the study. The definitions of each are:

CKD progression: the composite of a >50% decrease in eGFR or development of end-stage renal disease (ESRD) requiring chronic dialysis or kidney transplantation. For participants without CKD at baseline, CKD progression will be the composite of a >30% decrease in eGFR and an end value of<60 ml/min/1.73m2.

Incident albuminuria: a doubling of urinary albumin-to-creatinine (ACR) ratio from a value <10 mg/g to a value of >10 mg/g. This outcome applies to CKD and non-CKD subjects.

*Quality of life and other related measures -* Health-related quality of life (HRQoL) is measured by EuroQoL Group 5-Dimension Self-Report Questionnaire (EQ-5D); depression on the 9-item patient health questionnaire (PHQ-9); anxiety on the Zung self-rating anxiety scale (SAS); self-efficacy on the short fall self-efficacy scale international (FES-I); and frailty on the FRAIL scale.

*Major bleeding -* defined according to the International Society on Thrombosis and Haemostasis (ISTH) criteria: (i) fatal bleeding, and/or (ii) symptomatic bleeding in a critical area or organ, such as intracranial, intraspinal, intraocular, retroperitoneal, intraarticular or pericardial, or intramuscular with compartment syndrome, and/or; (iii) bleeding causing a fall in hemoglobin level of 20 g/L (1.24 mmol/L) or more, or leading to transfusion of two or more units of whole blood or red cells.

*PAD -* This includes systemic embolism, carotid and peripheral revascularization, abdominal aortic aneurysm repair, and other objectively defined PAD events

*Coronary artery revascularization -* This includes percutaneous coronary intervention (PCI) or coronary artery bypass grafting (CABG).

#### 2.5.3 Safety outcomes

All serious adverse events (SAEs) and adverse events of special interest (AESI) according to standard definitions will be collected until study end.

*Serious Adverse Event (SAE)* is consistent with NHLBI guidelines that meet any of the following criteria: fatal or life-threatening, result in significant or persistent disability, requires or prolongs hospitalization, results in a congenital anomaly/birth defect, or an important medical events that investigators judge to represent significant hazards or harm to research participants and may require medical or surgical intervention to prevent one of the other outcomes listed in this definition (e.g. hospitalization, death, persistent disability).

*Adverse Events (or symptoms) of Special Interest (AESI)* are expected adverse effects of intensive BP lowering that are of special interest are listed below. To better assess participants’ tolerability to the study treatments, the following AESIs and whether they are new or ongoing from baseline, will be reported in the eCRF at specified visits, regardless of severity and seriousness:

- *Hypotension:* SBP <90 mmHg with hypotensive symptoms (dizziness, etc.).
- *Orthostatic hypotension:* a drop in SBP of at least 20 mmHg or in DBP of at least 10 mmHg within 3 minute after the participant stands, as compared with the value obtained when the participant is seated;
- *Syncope:* a transient loss of consciousness with fall or near fall due to cerebral hypoperfusion, characterized by a rapid onset, short duration, and spontaneous complete recovery;
- *Bradycardia:* heart rate <40 bpm on ECG;
- *Electrolyte abnormality:* serum sodium ≤130 or >150 mEq/L or serum potassium <3.0 or >5.5 mEq/L;
- *Injurious fall:* a fall resulting in an evaluation in an emergency department or that resulted in hospitalization;
- *Acute kidney injury or acute renal failure:* is this is a coded diagnosis listed in the hospital discharge summary and is believed by the safety officer to be one of the top three reasons for admission or continued hospitalization.

## 3 Analysis principles

### 3.1 Sample size

The primary objective of the study is to determine whether more-intensive long-term BP control is superior to standard guideline-recommended BP control. This objective will be assessed using a hierarchical composite endpoint and analyzed using the win-ratio method. It was observed that the blinded pooled annual event rates for CV death, non-fatal stroke, non-fatal MI and HF hospitalization were 2%, 2%, 0.5%, and 4%, respectively, in the standard arm, during a blinded review of the grouped data in the study in 2023. The assumption is that intensive treatment will result in relative reductions of 43%, 20%, 17%, and 38% for CV death, non-fatal stroke, non-fatal MI, and HF hospitalization, respectively, as was shown in the Systolic Blood Pressure Intervention Trial (SPRINT).^2^ Based on a power of 80% and a 2-sided α of 5%, a win-ratio effect size of 1.50 (which is similar to that observed in SPRINT between the intensive group and standard group) is likely to be achieved with a sample size of 1624 participants followed up for a mean of 3 years. The goal is to enroll 1675 participants by December 2024, assuming 3% of participants are lost to follow-up.

### 3.2 Software

Analyses will be conducted primarily using SAS Enterprise Guide (version 8.3 or above) and R (version 4.0.0 or above).

### 3.3 Interim analyses

One formal interim analysis with win ratio was planned and undertaken. The Haybittle-Peto rule^3^ of an α <0.001 for the interim analysis in favor of intensive BP control will be applied to be considered significant to justify halting or modifying the study before the planned completion of recruitment. The α-level for the final analysis will be the conventional significance level (α = 0.05) given the infrequent interim analysis.

### 3.4 Multiplicity adjustment

Statistical tests are two-sided with a nominal level of 5%. For all secondary outcomes, the family-wise error rate will be controlled by applying a sequential Holm-Sidak correction.^4^ Briefly, the approach consists of ordering all p-values from smallest to largest, and then comparing them to an adjusted level of significance calculated as 1-(1-0.05)^1/C^, where C indicates the number of comparisons that remain. The sequential testing procedure stops as soon as a p value fails to reach the corrected significance level. This will apply only to the primary analysis of an outcome (i.e. not to sensitivity analyses). No multiplicity adjustment will be applied to safety outcomes.

### 3.5 Data sets analysed

#### 3.5.1 Analysis populations

The intention-to-treat (ITT) population: all patients who were randomized regardless of diagnosis and whether they received study treatment according to the protocol, and excluding those who withdrew their consent for any data to be used.

#### 3.5.2 Analysis strategy

The analysis will follow the ITT principle. Baseline characteristics will be summarized by treatment group. Continuous variables will be summarized by means (SD), and medians (IQR). The hierarchical primary outcome will be analysed by win-ratio analysis and each participant in the intensive group will be compared with each participant in the usual-care group to determine the win/loss/tie in each pair across each of the hierarchical outcomes. Secondary analysis will include conventional time-to-event outcomes using Cox proportional hazard models between the two treatment arms.

## 4 Planned analyses

### 4.1 Subject disposition

The flow of patients through the trial will be displayed in a Consolidated Standards of Reporting Trials (CONSORT) diagram. The report will include: the number of screened patients who met study inclusion criteria and the number of patients included; and reasons for exclusion of non-included patients.

### 4.2 Patient characteristics and baseline comparisons

Baseline characteristics will be summarized by treatment groups. Discrete variables will be summarised by frequencies and percentages. Percentages will be calculated according to the number of patients for whom data are available. Continuous variables will be summarized using mean and SD, and median and interquartile range (Q1-Q3). No statistical test will be performed on baseline characteristics.

### 4.3 Analysis of the primary outcome

The primary outcome of the trial is a hierarchical composite of time to CV death, number of strokes, time to the first stroke, number of MI events, time to the first MI, number of HF hospitalization events, and time to the first HF hospitalization. The hierarchical composite primary outcome will be analyzed by the win-ratio approach.

#### 4.3.1 Main analysis

The main analysis will be performed in the ITT population using the win-ratio approach.^5^

Treatment allocation (intensive BP control vs standard BP control) will be included as groups to be compared. The effect of the intervention will be presented as the win ratio of intensive BP control relative to standard BP control, and its 95% confidence intervals (CI). A win ratio greater than 1 corresponds to a better hierarchical primary outcome for intensive BP control compare to standard BP control. In addition, win odds and net benefit will also be reported.

To perform the unmatched win ratio analytical approach, each participant in the intensive BP control group is paired with each participant in the standard BP control group, forming *n*_l_ × *n*_2_ pairs, where *n*_l_ is the total number of participants in the intensive BP control group and *n*_2_ is the total number of participants in the standard BP control group. These *n*_l_ × *n*_2_ pairs are then classified into outcomes categories (a) – (o) (defined in Table 1). Note that categories (a) and (b) take priority over (c) and (d): any pair is only classified as having greater number of strokes if it is not known who had CV death first. The same logic applies to all outcome categories in Table 1.

**Table 1.** Categories for the hierarchical composite primary outcome.

|  |  |
| --- | --- |
| (a) | Intensive BP control patient has CV death first |
| (b) | Standard BP control patient has CV death first |
| (c) | Intensive BP control patient has greater number of strokes |
| (d) | Standard BP control patient has greater number of strokes |
| (e) | Intensive BP control patient has the first stroke event first |
| (f) | Standard BP control patient has the first stroke event first |
| (g) | Intensive BP control patient has greater number of MI events |
| (h) | Standard BP control patient has greater number of MI events |
| (i) | Intensive BP control patient has the first MI event first |
| (j) | Standard BP control patient has the first MI event first |
| (k) | Intensive BP control patient has greater number of HF hospitalization events |
| (l) | Standard BP control patient has greater number of HF hospitalization events |
| (m) | Intensive BP control patient has the first HF hospitalization event first |
| (n) | Standard BP control patient has the first HF hospitalization event first |
| (o) | None of the above. |

Let *N_a_* to *N_o_* denote the number of pairs in outcome categories a to o, and then 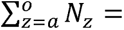 *n*_l_ x *n*_2_. Categories b, d, f, h, j, l and n are winners for the intensive BP control group, with *N_b_* + *N_d_* + *N_f_* + *N_h_* + *N_j_* + *N_l_* = *N_Win_*, while categories a, c, e, g, i, k and m are losers, with *N_a_* + *N* + *N_e_* + *N_g_* + *N_i_* + *N* = *N_lOSe_*. The win ratio can be calculated as *R_W_* = *^N^Win^⁄N^lOSe*, win odds can be calculated similarly as *Odds_W_* = (*N_Win_* + 0.5 • (*n*_l_ x *n*_2_-*N*_Win_ -*N*_lOSe_))⁄(*N*_lOSe_ + 0.5 • (*n*_l_ x *n*_2_ -*N*_Win_ -*N*_lOSe_ )), net benefit as *NB* = (*N_Win_ -N_lOSe_*)/ (*n*_l_ x *n*_2_).

To obtain the p value of the win ratio, we compare all possible pairs of patients *i, J* to determine whether patient *i* was the winner, the loser or they tied irrespective of treatment group. Assignment of *u_ij_* = +1,-1 or 0 will be applied according to whether patient *i* was the winner, the loser or they tied. Then, for patient *i*, to define *u_i_* = ∑*_i=j_ u_ij_*. Note *u_i_* will be a positive integer if patient *i* wins more often than he loses compared to all other patients. Then calculate 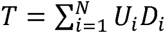, where *D_i_* =1 if patient *i* is on intensive BP control and *D_i_* =0 if patient *i* is on standard BP control. Under the null hypothesis of no true difference between intensive and standard BP control, *r* has variance *v* where 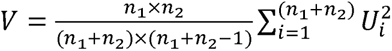. Then 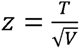is a standardized normal deviate from which the p value can be readily obtained. Specifically, *z* > 1.96, *z* > 2.38 and *z*> 3.29 correspond to P<0.05, P<0.01 and P<0.001, respectively.

In addition, to obtain the 95%CI of the win ratio, under the null hypothesis, the log win ratio is approximately normally distributed and its standard error can be estimated by 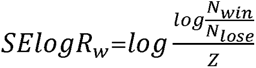, where Z is the Z-statistic obtained from the Finkelstein-Schoenfeld test. A 95% confidence interval can then be calculated for the log win ratio and this can be back-transformed to give a 95% confidence interval for the win ratio, i.e., 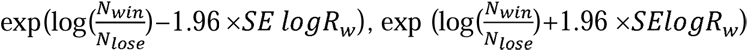.

Results will be presented by an overall win ratio for the primary hierarchical composite endpoint and a cumulative win ratio for each component of the hierarchical composite endpoint. ^6^ See an example in Figure 1 below.

**Figure 1.**
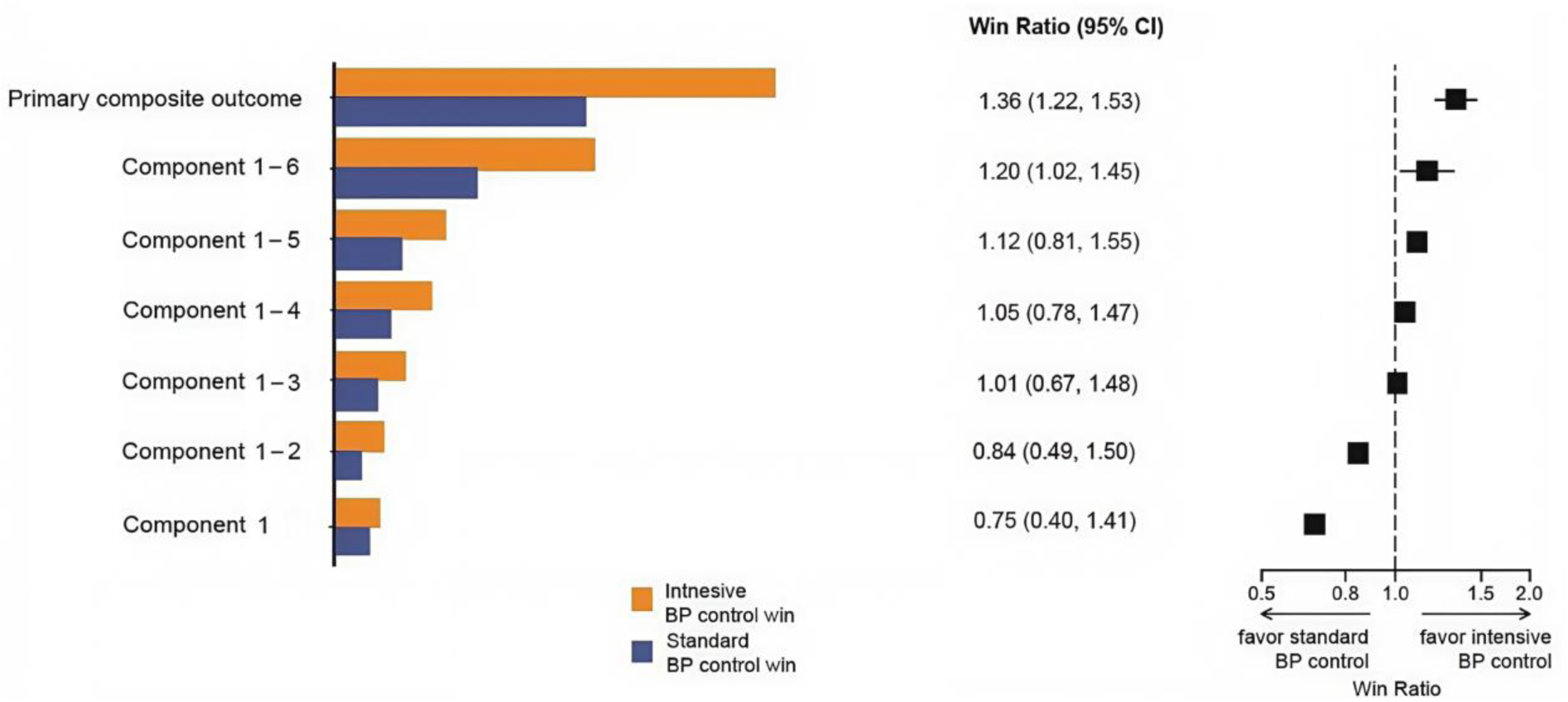
Win ratio for overall and cumulative win ratio for each component of the primary composite endpoint.

#### 4.3.2 Subgroup analyses

Seven pre-specified subgroup analyses will be carried out, irrespective of whether there is a significant treatment effect on the primary outcome. These analyses will only be performed in the ITT population.

Subgroups based on patient characteristics assessed before randomisation are defined as follows:

- baseline BP (<130 vs. ≥130 mmHg);
- sex (male vs female);
- age (<75 vs. ≥75 years);
- history of CKD;
- history of ASCVD;
- anticoagulation use;
- CHA_2_DS_2_-VASc score (0-1 vs. ≥2).

where the definition of ASCVD is:

1) Prior history of thromboembolism: defined as any of the following criteria: a) Ischemic stroke; b) Transient ischemic attack (TIA); c) Systemic embolism (SE);
2) Coronary artery disease or peripheral artery disease: defined as any of the following criteria:

a) Previous myocardial infarction (MI), percutaneous coronary intervention (PCI), coronary artery bypass grafting (CABG), carotid endarterectomy (CE), carotid stenting; b) Peripheral artery disease (PAD) with revascularization; c) Acute coronary syndrome, positive exercise stress test, or positive cardiac imaging examination; d) At least 50% narrowing in the lumen diameter of coronary arteries, carotid arteries, or lower limb arteries; e) Abdominal aortic aneurysm with a diameter ≥5cm, regardless of whether repair surgery has been performed.

The analysis for each subgroup will be performed using win ratio stratified for each subgroup. Within each subgroup, summary measures will include number of participants in each treatment arm and win ratio for the treatment effect with 95% CI. The results will be displayed on a forest plot with the win ratio estimate for the treatment arm in each subgroup.

#### 4.3.3 Treatment of missing data

No imputation will be conducted.

#### 4.3.4 Sensitivity analyses

The primary analyses of the hierarchical composite outcome will be repeated using matched win ratio analysis, where participants in the intensive and standard BP control groups are first matched into pairs by the the components of the CHA_2_DS_2_-VASc history of Congestive heart failure, history of Hypertension, Age ≥75 years or nothistory of Diabetes mellitus, history of Stroke or TIA or thromboembolism, history of Vascular disease [includes which includes PAD, MI, or aortic atheroma]) using nearest neighbour matching method, instead of using all possible pairs between the intensive and standard BP control groups. For each matched pair, one examines the outcome in each category in Table 1 and then calculate the overall win ratio similar to unmatched pair approach. Denote 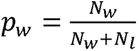, and the win ratio 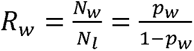. The 95% CI of *p_W_* can be calculated as

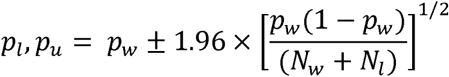

It follows that the 95% CI for *R_W_* is 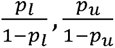. For significance test, l/2 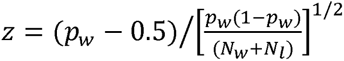 is a standard normal deviate under the null hypothesis, readily yielding the required P value.

Time to the composite outcome of CV death, stroke, MI or HF hospitalization will also be analysed using Cox proportional hazard models between intensive and standard BP control groups.

### 4.4 Analysis of secondary outcomes

All secondary outcome analyses will be performed in the ITT population.

#### 4.4.1 Each component of the primary outcome

Time to CV death, time to the first stroke, time to the first MI and time to the first HF hospitalization will be analyzed using Cox proportional hazard models between intensive and standard BP control groups. These time to events outcomes are included as the dependent variable, and treatment allocation is included as binary independent variable. To account for the stratification variables and to maximise precision, baseline BP, sex, age, history of ASCVD, will also be included as covariates. The effect of intensive BP control relative to standard BP control will be presented as HR and its 95%CI, where an HR greater than 1 corresponds to an increase of risk in the corresponding events in the intensive BP control compared to the standard BP control group. We will also calculate number need to treat (NNT) and number needed to harm (NNH) using survival analysis.

In addition, number of strokes, number of MI events and number of HF hospitalization events, will be analyzed using Poisson log-linear regression model between intensive and standard BP control groups with length of follow-up included as an ‘offset’ in the model. These count outcomes are included as the dependent variable, and treatment allocation is included as binary independent vatiable. To account for the stratification variables and to maximize precision, baseline BP (above or below median), sex (male vs female), age (<75 vs ≥75 years), history of ASCVD, will also be included as covariates. The effect of intensive BP control relative to standard BP control will be presented as beta and its 95%CI, where a beta greater than 0 corresponds to an increase of risk in the corresponding number of events in the intensive BP control compared to the standard BP control group.

#### 4.4.2 All-cause death

Time to all-cause death is analysed using Cox proportional hazard models between intensive and standard BP control groups. Time to all-cause death is included as the dependent variable, and treatment allocation is included as binary independent variable. To account for the stratification variables and to maximize precision, baseline BP (above or below median), sex (male vs female), age (<75 vs ≥75 years), history of ASCVD, will also be included as covariates. The effect of intensive BP control relative to standard BP control will be presented as HR and its 95%CI, where an HR greater than 1 corresponds to an increase of all-cause death risk in the intensive BP control compared to the standard BP control group.

#### 4.4.3 Time to first occurrence of progression of CKD or incident albuminuria

Time to first occurrence of progression of CKD or incident albuminuria is analysed using Cox proportional hazard models between intensive and standard BP control groups. Time to first occurrence of progression of CKD (defined by decline in creatinine and according to macro- and micro-albuminuria, as in the SPRINT trial) or incident albuminuria is included as the dependent variable, and treatment allocation is included as binary independent variable.

To account for the stratification variables and to maximize precision, baseline BP (above or below median), sex (male vs female), age (<75 vs ≥75 years), history of ASCVD, will also be included as covariates. The effect of intensive BP control relative to standard BP control will be presented as HR and its 95%CI, where an HR greater than 1 corresponds to an increase of risk for first occurrence of progression of CKD or incident albuminuria in the intensive BP control compared to the standard BP control group.

#### 4.4.4 HRQoL

Each component domain of the EQ-5D-3L (difference between 12-month and baseline) scale will be analysed via ordinal logistic regression model. The visual analogous scale (score of 0 to 100) will be analysed using the same approach but with linear regression (i.e. assuming a normal distribution and an identity link function). The overall health utility EQ-5D-3L score will be calculated using Chinese weights and compared between groups in a similar manner to that used for the visual analogous scale. We do not plan to undertake adjusted or subgroup analyses.

#### 4.4.5 Major bleeding according to the ISTH criteria

Major bleeding will be analyzed using binary logistic regression model, with major bleeding events (0 or 1) as the dependent variable and treatment allocation as the independent variable, with length of follow up included as an off set in the model. To account for the stratification variables and to maximize precision, baseline BP (above or below median), sex (male vs female), age (<75 vs ≥75 years), history of ASCVD, will also be included as covariates. The effect of intensive BP control relative to standard BP control will be presented as odds ratio (OR) and its 95% CI, where an OR greater than 1 corresponds to an increase of risk in major bleeding in the intensive BP control compared to the standard BP control group.

#### 4.4.6 PAD

PAD will be analyzed using binary logistic regression model, with PAD event (0 or 1) as the dependent variable and treatment allocation as the independent variable, with length of follow up included as an off set in the model. To account for the stratification variables and to maximize precision, baseline BP (above or below median), sex (male vs female), age (<75 vs ≥75 years), history of ASCVD, will also be included as covariates. The effect of intensive BP control relative to standard BP control will be presented as OR and its 95% CI, where an OR greater than 1 corresponds to an increase of PAD in the intensive BP control compared to the standard BP control group.

#### 4.4.7 Coronary artery revascularization

Time to coronary artery revascularization is analysed using Cox proportional hazard models between intensive and standard BP control groups. Time to coronary artery revascularization is included as the dependent variable, and treatment allocation is included as binary independent variable. To account for the stratification variables and to maximize precision, baseline BP (above or below median), sex (male vs female), age (<75 vs ≥75 years), history of ASCVD will also be included as covariates. The effect of intensive BP control relative to standard BP control will be presented as HR and its 95% CI, where an HR greater than 1 corresponds to an increase of risk in coronary artery revascularization in the intensive BP control compared to the standard BP control group.

### 4.5 Analysis of SAEs and AESI

SAEs will be summarised as the number of events as well as the number and proportion of patients experiencing at least one SAE/AESI event. This will be done overall and by category of event according to Medical Dictionary for Regulatory Activities (MeDRA) system organ classes and preferred terms. The overall proportion of patients with SAE/AESIs in the intensive and standard BP control will be described. Primary and underlying causes of deaths will be summarised by treatment arm with no formal test. A listing of all SAE/AESIs will be compiled (in an appendix).

## Data Availability

All data produced in the present study are available upon reasonable request to the authors

## 6 Appendix 1: Proposed Tables and figures

### 6.1 Tables

**Table 1.**
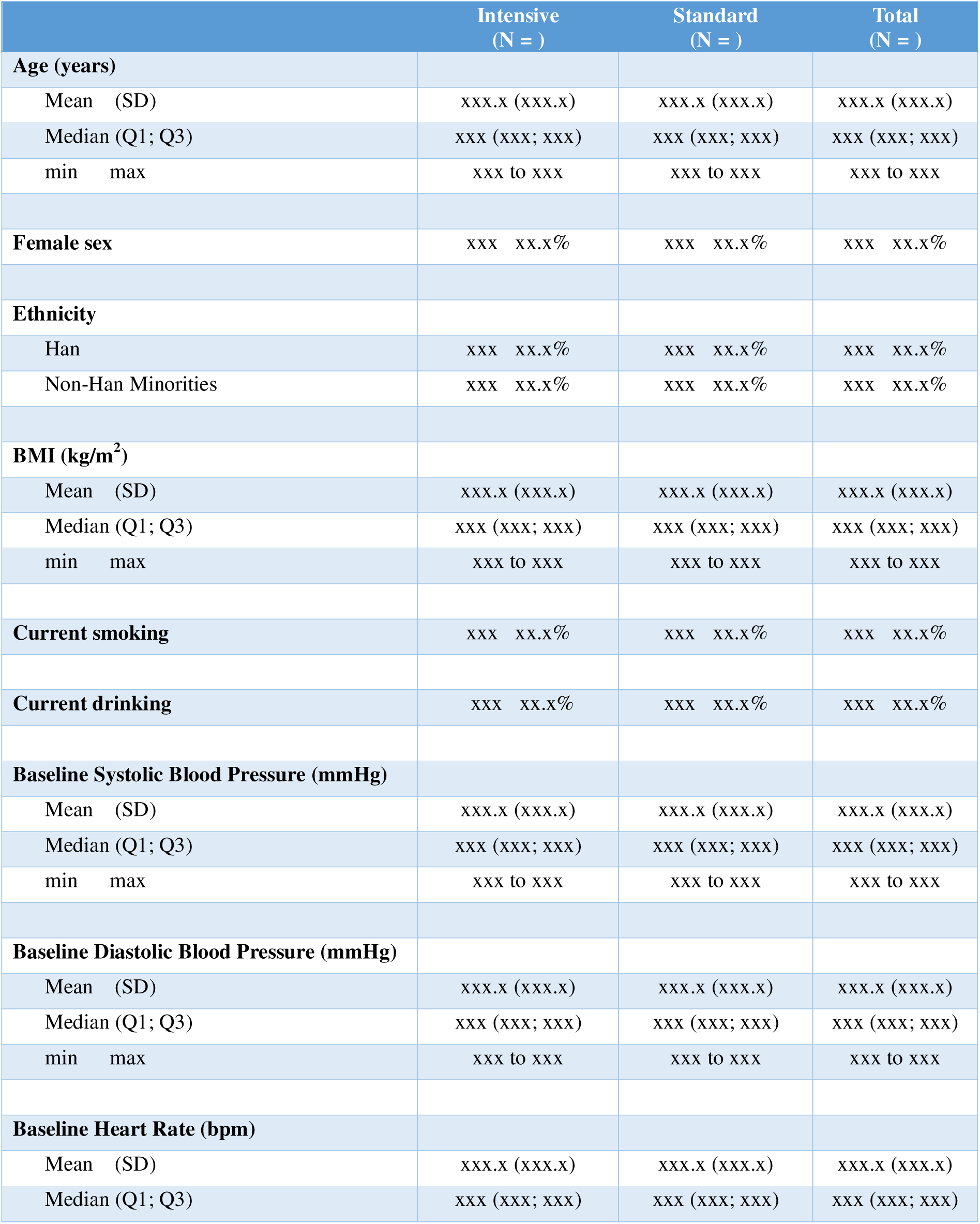
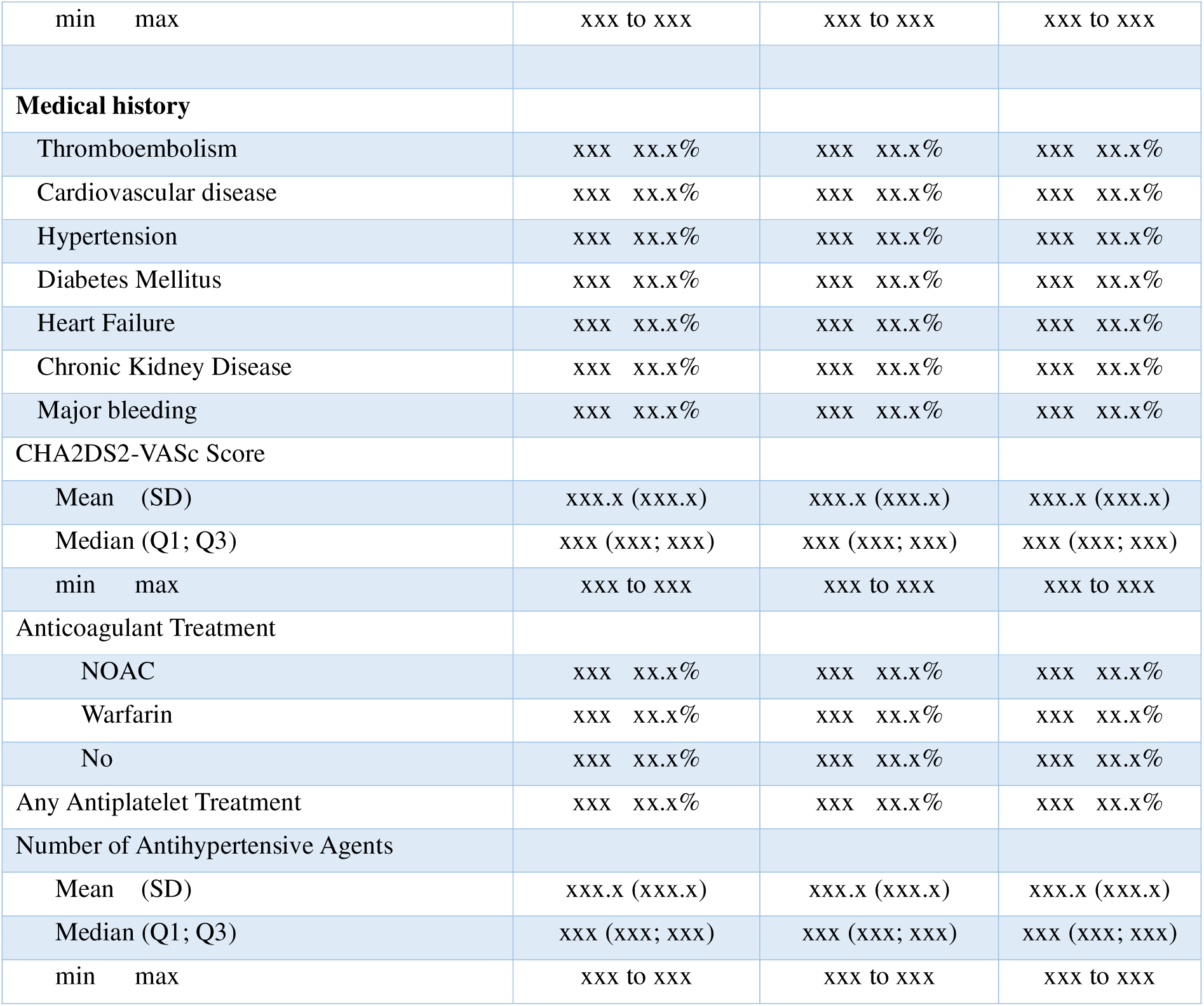
Baseline characteristics. **Definition:** Thromboembolism: TIA/Ischemic Stroke/ Systemic Embolism; Vascular disease: Coronary Heart Disease/ Peripheral Arterial Disease/ Abdominal Aortic Aneurysm; Chronic Kidney Disease: baseline estimated glomerular filtration rate <60 ml /min/1.73 m^2^

**Table 2.**
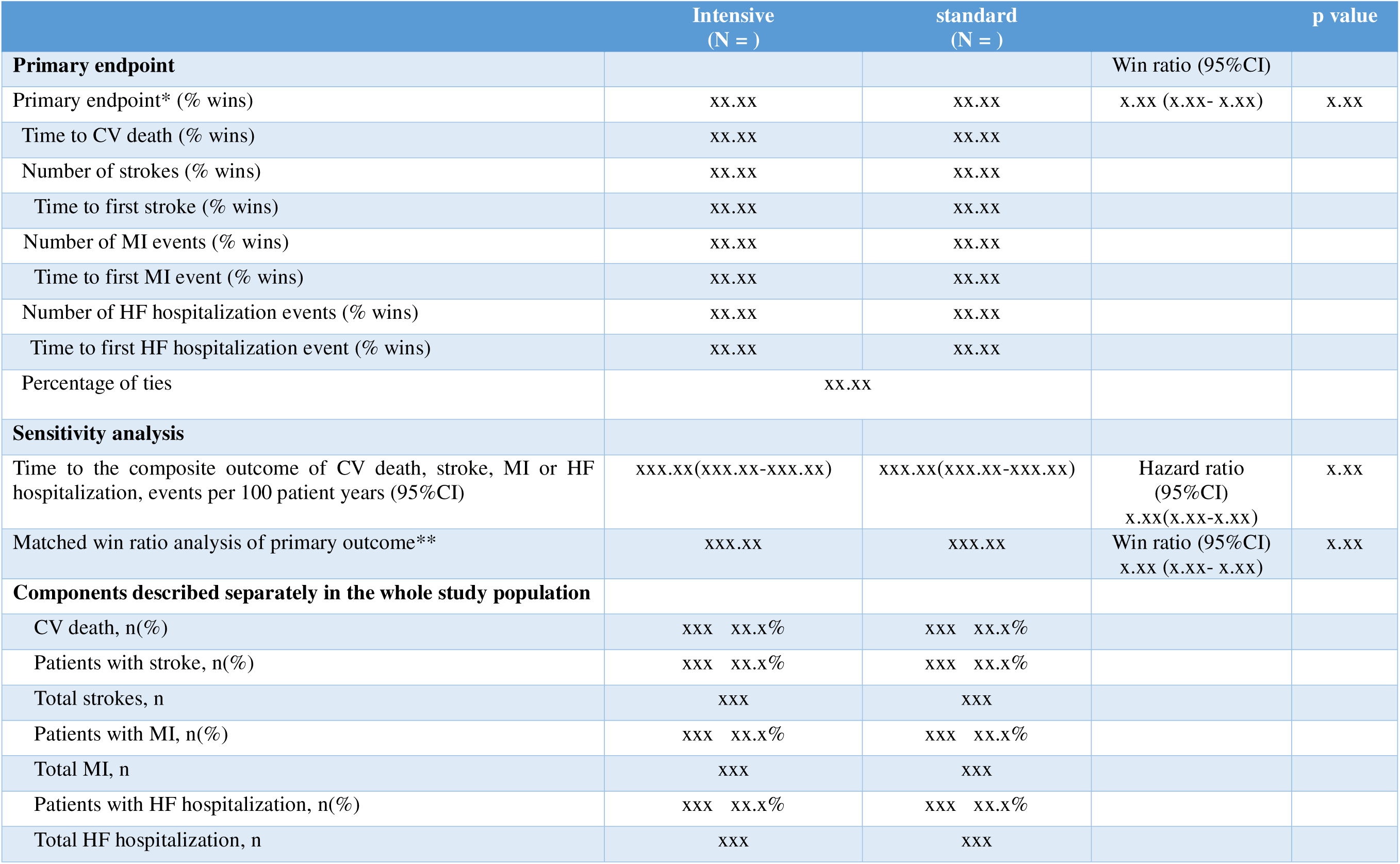
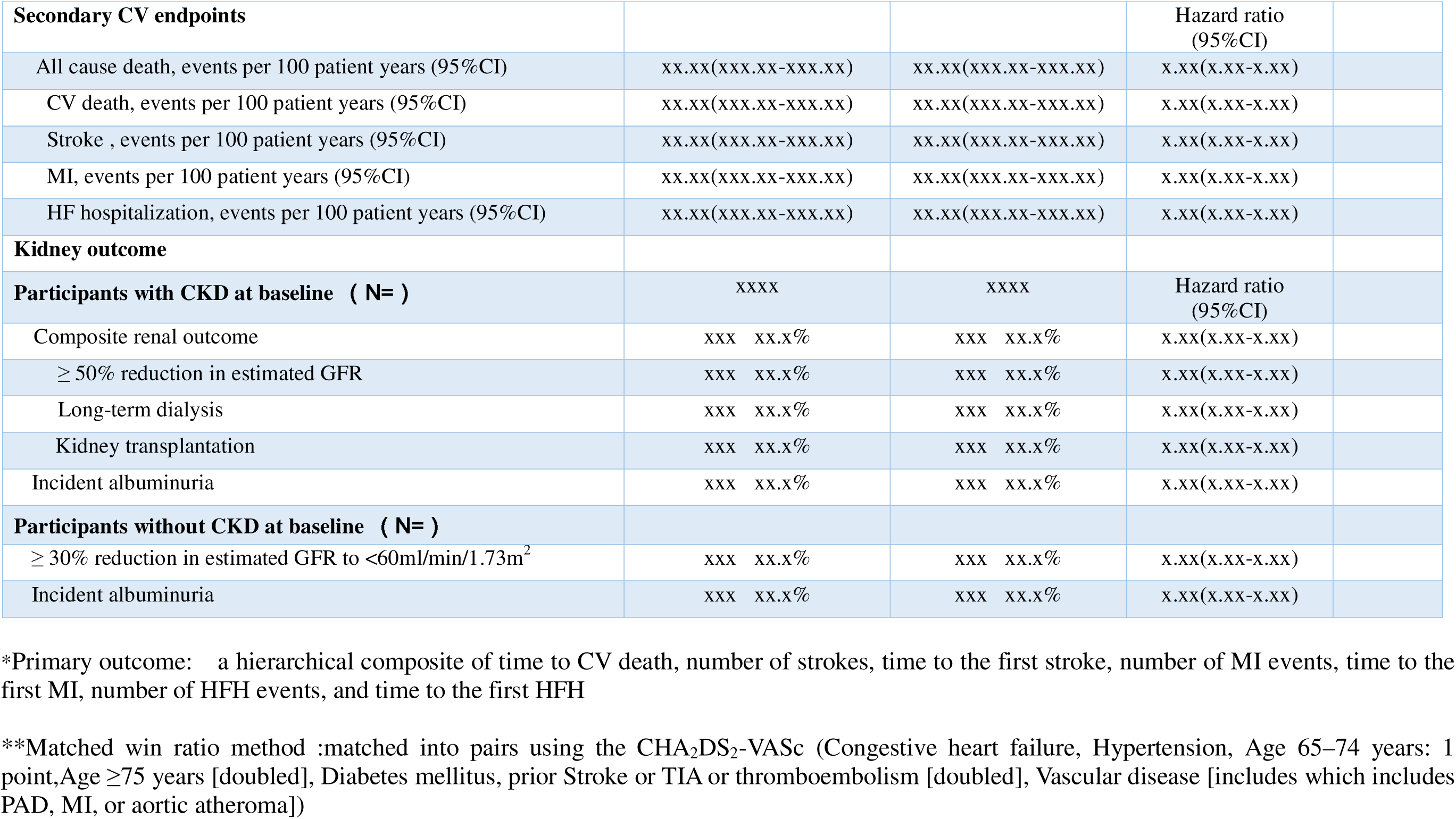
Descriptive table of primary outcome and other secondary outcomes.

**Table 3.** Descriptive table of SAEs and AESIs.

| Variable | Intensive<br>(N = ) | Standard<br>(N = ) |
| --- | --- | --- |
|  | Number of patients(%) |  |
| **Serious adverse event** | xxx(xx.x) | xxx(xx.x) |
| **Conditions of interest** |  |  |
| Serious adverse event only |  |  |
| Hypotension | xxx (xx.x) | xxx (xx.x) |
| Syncope | xxx (xx.x) | xxx (xx.x) |
| Bradycardia | xxx (xx.x) | xxx (xx.x) |
| Electrolyte abnormality | xxx (xx.x) | xxx (xx.x) |
| Injurious fall | xxx (xx.x) | xxx (xx.x) |
| Acute kidney injury or<br>acute renal failure | xxx (xx.x) | xxx (xx.x) |
| **Monitored clinical events** |  |  |
| Serum sodium <130<br>mmol/liter | xxx.x (xxx.x) | xxx.x (xxx.x) |
| Serum sodium >150<br>mmol/liter | xxx.x (xxx.x) | xxx.x (xxx.x) |
| Serum potassium <3.0<br>mmol/liter | xxx.x (xxx.x) | xxx.x (xxx.x) |
| Serum potassium >5.5<br>mmol/liter | xxx.x (xxx.x) | xxx.x (xxx.x) |
| Orthostatic hypotension | xxx.x (xxx.x) | xxx.x (xxx.x) |

### 6.2 Figures

**Figure 1:**
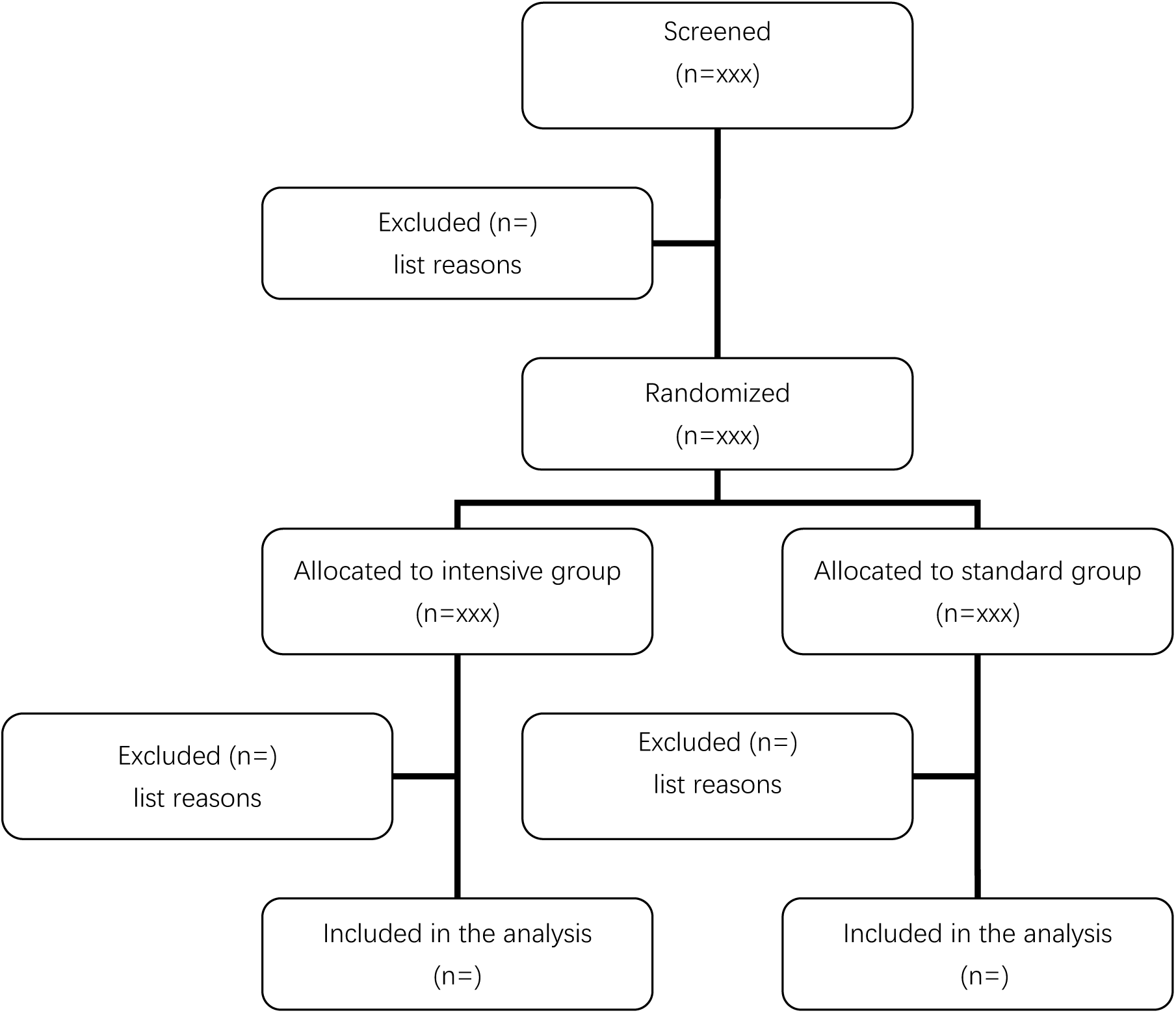
Consort flowchart.

**Figure 2.**
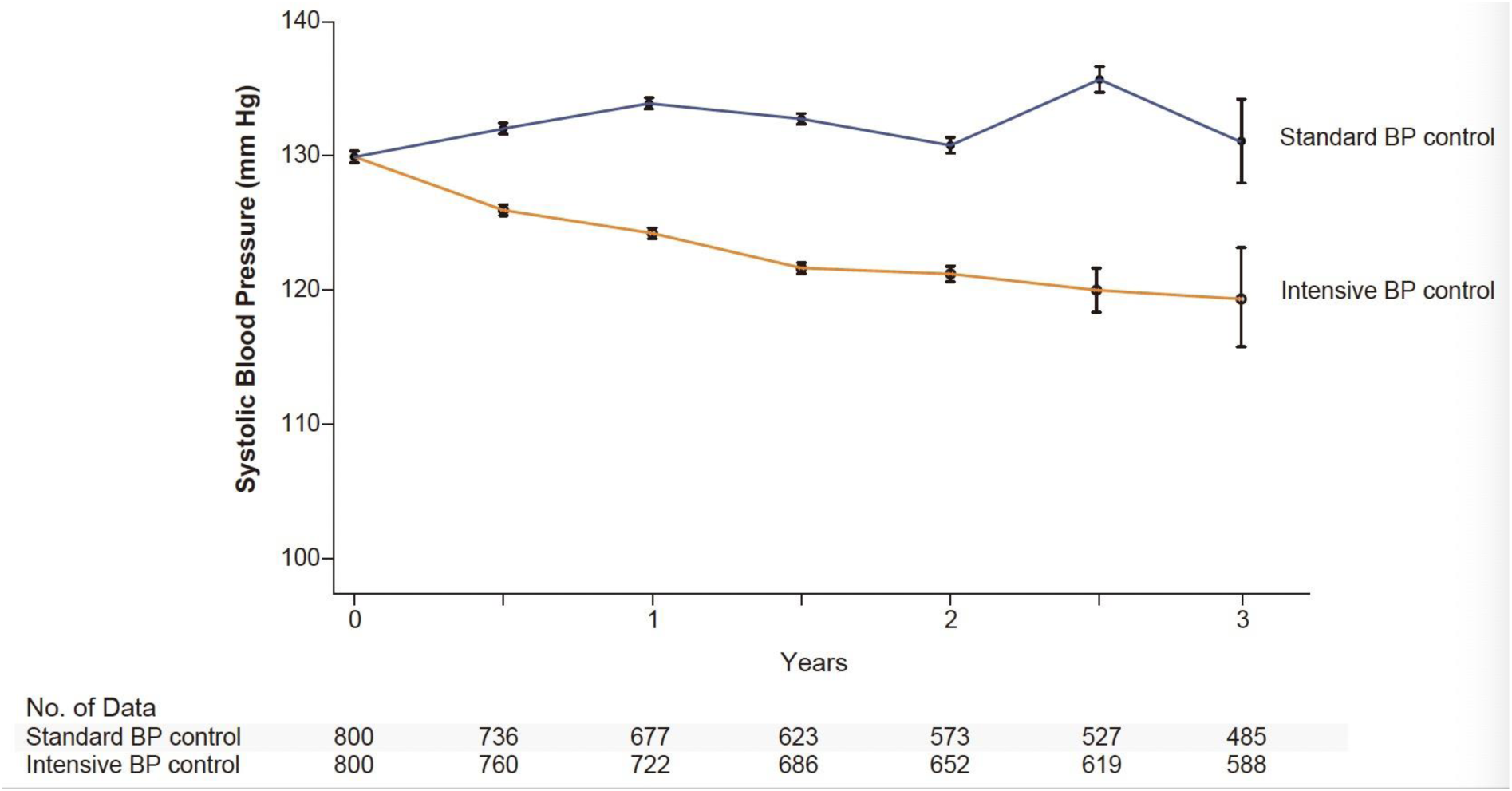
Blood pressure in the two groups (for example)

**Figure 3.**
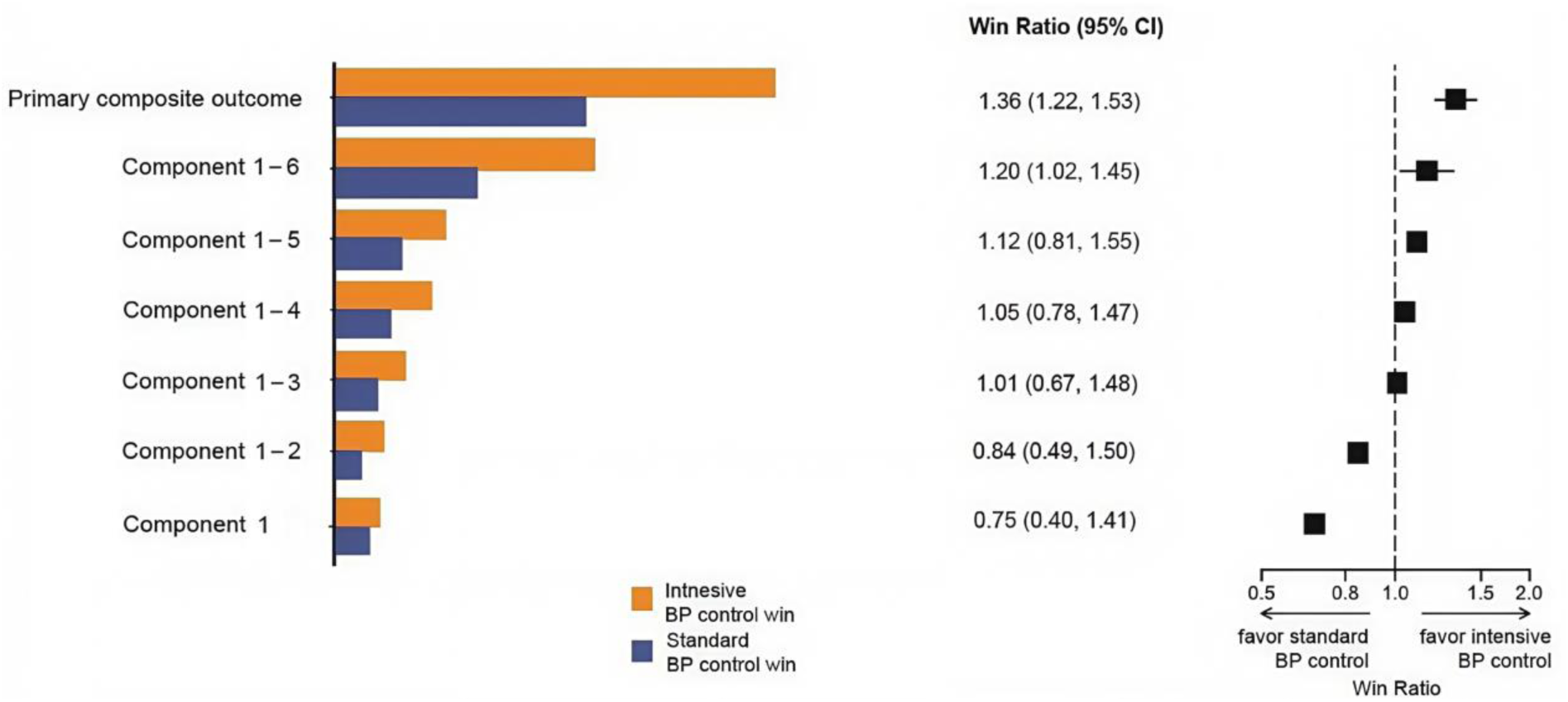
win-ratio bars of the primary outcome (for example)

**Figure 4.**
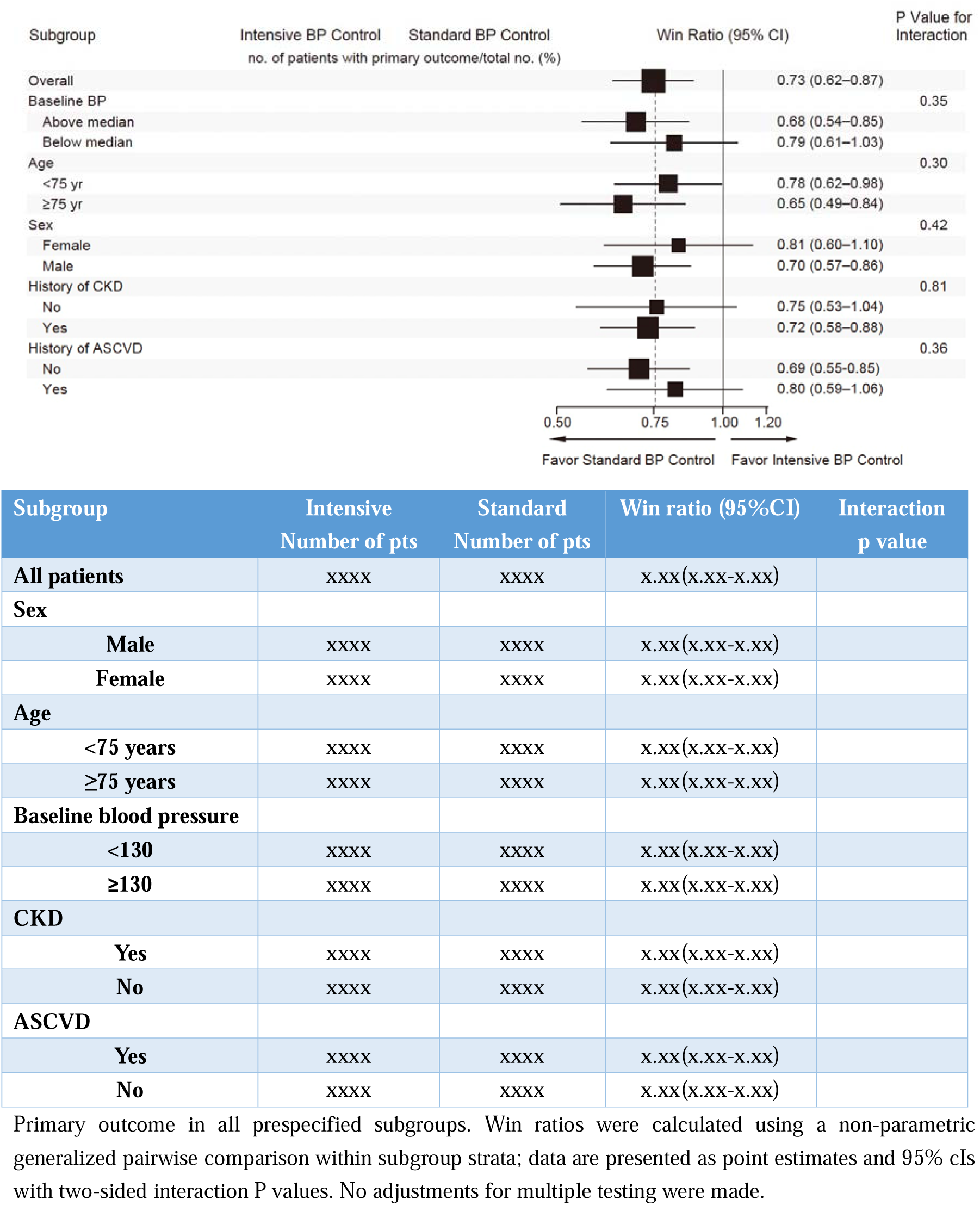
Primary outcome in all prespecified subgroups(for example)

